# Drug Resistant Profile Among TB Patients From Haut-Ogooué Region Gabon

**DOI:** 10.64898/2026.01.06.26343446

**Authors:** Jabar Babatunde Pacome Agbo Achimi Abdul, Paul Snael Eyinghi Openbindjala, Micheska Epola Dibamba Ndanga, Saïdou Mahmoudou, Stredice Manguiga, Tiburce Etele, Arnault Guy Rogue Ibinda Mfoumbi, Audrey Sylviane Imounga, Guy Roger Ndong Atome, Joseph Privat Ondo, Louis Clément Obame Engonga, Cedric Obiang Sima

**Affiliations:** Centre de Recherches Médicales de Lambarene, Laboratoire National de Reference pour la Tuberculose, Lambarene, Gabon; Laboratoire de Substances Naturelles et de Synthèses Organométalliques (LASNSOM), Université des Sciences et Techniques de Masuku, Franceville, Gabon; Laboratoire de Recherche en Biochimie (LAREBIO), Faculté des Sciences, Université des Sciences et Techniques de Masuku, Franceville, Gabon; Programme National de Lutte contre la Tuberculose, Libreville, Gabon; Centre Hospitalier Universitaire Amissa Bongo de Franceville, Franceville, Gabon; Hopital Specialise de Nkembo, Libreville Gabon

**Author notes:** **Corresponding Author** : Jabar Babatundé Pacome Achimi Agbo Abdul.

**Keywords:** tuberculosis, TB, drug resistance, HIV

## Abstract

Drug-resistant tuberculosis (DR-TB) remains a major health concern, especially in high-burden areas like Gabon’s Haut-Ogooué region. This study investigates the prevalence of drug-resistant tuberculosis (TB) and associated factors in the Haut-Ogooué region. Drug resistance poses a significant threat to global TB control efforts. We conducted a retrospective and prospective study at Centre Hospitalier Universitaire de Franceville from August 2022 to August 2024. A total of 1,792 participants were included. Sputum samples were collected and analyzed using GeneXpert MTB/RIF ULTRA and MTB/XDR assays. Data on demographic characteristics, HIV status, and previous TB treatment history were collected and analyzed using R software. The majority of participants were male (62.1%). Multidrug-resistant TB (MDR-TB) was the predominant resistance profile (94.44% of resistant isolates). A significant association was found between previous TB treatment history and drug-resistant TB (p < 0.001). No significant associations were observed between drug-resistant TB and sex or age. A high proportion of participants had unknown HIV status (47.8%), and most resided in urban areas (85.5%). The high prevalence of MDR-TB and its association with previous TB treatment highlight the urgent need to strengthen TB control programs, improve treatment practices, and address the social determinants of TB in Gabon.

## Introduction

Tuberculosis (TB) caused by the Mycobacterium tuberculosis complex is still a major global health problem. Although diagnosis and treatment for TB have improved, TB remains a leading death from an infectious disease. According to the World Health Organization (WHO), in 2022 10.6 million people were ill with TB and 1.3 million died [1]. It also includes 167,000 deaths among people living with HIV (PLHIV). Drug resistant TB strains emerging and spreading is a formidable challenge to global TB control efforts and a risk to derail end TB strategy goals set by WHO [1]. The World Health Assembly adopted the End TB Strategy in 2014 to end the global TB epidemic with ambitious targets for reducing TB incidence by 90% and TB deaths by 95% by 2035 as compared to 2015 levels. Achieving these targets is a major fear in efforts to overcome this obstacle (drug resistance) [2].

MDR TB (multidrug resistant TB) is TB that is resistant to at least isoniazid and rifampicin (the most powerfull first line anti TB drugs) [1]. According to the WHO, it is estimated that in 2022, 410,000 cases of MDR/RR TB were encountered and this is a serious public health issue which can reiterate the progress being made in TB control [1]. Treatment of MDR-TB is more complex, longer and more expensive than for drug susceptible TB, with lower success rates, higher mortality [3]. In addition, second line anti-TB drugs, used to treat MDR-TB, are in general more toxic and less effective than first line drugs and so patient management is further complicated with increased risk of adverse drug reaction [4]. Prolonged treatment duration for MDR-TB is 18 months or more, and this has a huge impact on patients and the health system as such treatment often fails and patients do not adhere to treatment and may develop drug resistance further [5].

In Sub-Saharan Africa the burden of TB is particularly high, as this region faces the confluence of a number of factors that undermine the response to the epidemic. These include a high prevalence of HIV co-infection, which weakens the immune system and increases susceptibility to TB, and accelerates the progression from latent TB infection to active disease. In 2022 there were 23 percent of TB cases in the world in the WHO African Region [1]. Added to this is the growing proportion of TB in the continent that is highly resistant to drugs, caused by factors including poor treatment adherence, lack of control of essential anti TB medicines supply through stock outs, poor infection control measures in healthcare facilities which facilitates transmission of drug resistant strains [6].

Among the 30 high burden TB countries, Gabon is. According to WHO’s 2024 global TB report, Gabon has an incidence of 505 TB per 100,000 population[1]. Like many other resource limited settings, it is facing the challenge of rising rates of MDR-TB. In addition, co-infection rates for HIV in Gabon contribute to the TB burden due to the prevalence of HIV in the country[1]. HIV and TB interact bidirectionally as HIV increases the risk of TB disease and TB increases the risk of disease progression associated with HIV. However, while Gabon has a high burden of TB, drug resistance does raise growing concerns. With regard to the load of TB cases, Haut-Ogooué is one of the most populous regions of Gabon. Wildlife involvement and population movement in Haut Ogooué may affect both the transmission and drug resistance patterns of TB. Also, the region’s socioeconomic characteristics such as poverty levels, access to healthcare, may influence the TB epidemiology and treatment outcome. The gap in knowledge of the drug resistance profile of the TB patients in the Haut-Ogooué region is addressed by this study through the description of drug resistance pattern in the TB patients in the Haut-Ogooué region and the possible implication in the TB treatment and control strategies in Gabon

## Methods

### Ethical Approval

This study was approved by Centre Hospitalier Universitaire (CHR) de Franceville Institutional Ethics Committee (CEI) with the number CEI-023/2024FR. It guaranteed that the research was a research conducted within ethical principles and guidelines and the welfare of the individuals. All volunteers written informed consent was obtained before study enrolment for prospective sampling, the agreement to use the pre-existing samples was obtained from the CEI.

### Study design

We present the results of an combined retrospective and prospective study done between August 2022 and August 2024 at the Centre Hospitalier Universitaire de Franceville. This widened our dataset with both the known and unknown historical TB data to help understand the TB drug resistance profile in the region. Since it is a tertiary referral hospital in the Haut-Ogooué region, and since Franceville is going to be undergoing significant traffic flow alterations, including the reorganization of a round intersection, the Centre Hospitalier Universitaire de Franceville is a good site for this study. Two years duration of the study enabled large enough sample size to be recruited and data is collected over reasonable time period which might vary TB drug resistance patterns.

### Data Collection

Data regarding demographic and clinical information of participants were collected with a structured questionnaire and medical record review. The questionnaire yielded standardized information on other key variables such as sex, age, HIV status, and previous TB treatment history—variables known as risk factors for TB and drug resistance. Additional clinical details, as drawn from medical records and obtained as such, including prior TB treatment regimens and outcomes resulted from reviewing medical records further. The trained personnel guided data collection using appropriate data abstraction forms to ensure accuracy and consistency. The entire data including laboratory results and corresponding drug resistance profiles were tabulated in an Excel database.

### Sample Collection

The national guideline requires each participant to give two sputum samples. Mycobacterium tuberculosis is mainly cultured from sputum during the diagnosis of TB and is a important in the collection of the sputum samples. A spot sputum sample was obtained from one sample at the time of the initial patient visit. The second sample was an early morning sputum sample which is known to contain higher concentration of bacilli because the bacilli had accumulated overnight in the lungs. Patients were taught the proper technique for collection of sputum to help produce good quality samples. Temperature was controlled throughout transport of samples to the laboratory in a cool box into single use containers. Since the survival of Mycobacterium tuberculosis organisms depend on temperature, the use of temperature-controlled transport kept them alive, and the use of single use containers minimizes the risk of contamination. Samples were carried to the laboratory in a time frame, to maintain sample integrity and the accuracy of the laboratory tests.

### Laboratory Tests

The Genexpert MTB/Rif ULTRA assay, a rapid molecular test, detected Mycobacterium tuberculosis complex DNA and rifampicin resistance in all samples. Trained technicians followed the manufacturer’s protocol, ensuring result accuracy through quality control. Its high sensitivity, specificity, speed, and ease of use make it vital for quick TB diagnosis and rifampicin resistance detection.

The GeneXpert MTB/XDR assay rapidly identified isoniazid and multiple second-line TB drug resistance. Following standard procedures, sputum samples were liquefied using a 2:1 reagent-to-sample ratio with a 15-minute incubation. Results for TB detection and susceptibility to fluoroquinolones, isoniazid, and injectable drugs were available within 90 minutes, enabling timely treatment, especially crucial given increasing drug resistance.

### Data Analysis

Data analysis utilized R version 4.0.1. Descriptive statistics summarized demographics (frequencies/percentages for categorical variables like sex and HIV status; means/SD or medians/IQR for continuous variables like age). Resistance rates for individual and combined drugs were determined using proportions and 95% confidence intervals. Chi-square tests compared subgroups (e.g., HIV status, prior TB treatment), with p<0.05 indicating statistical significance.

## Results

A total of 1,792 participants were included in the study during the specified period. The baseline demographic and clinical characteristics of the study population are summarized in Table 1. The male gender constituted the majority of our study participants, accounting for 62.1% (1113/1792), while females comprised 37.9% (679/1792). This male predominance is a consistent finding in TB epidemiology worldwide. Various factors may contribute to this gender disparity, including differences in health-seeking behaviors, occupational exposures, biological susceptibility, and socio-cultural influences affecting disease reporting and healthcare access. Recognizing this imbalance is essential for tailoring public health interventions to ensure equitable TB control strategies that effectively address both genders. Table 1 shows baseline characteristics of research participants.

**Table 1.** Baseline Characteristics of study population (N = 1792)

| Variable |  | n | % | 95%CI |
| --- | --- | --- | --- | --- |
| Sex | Female | 679 | 37.9 | 35.6 - 40.2 |
|  | Male | 1,113 | 62.1 | 59.8 - 64.4 |
| Age (Years) | [0-24] | 489 | 27.3 | 25.2 - 29.4 |
|  | [25-44] | 677 | 37.8 | 35.5 - 40.1 |
|  | [45+] | 526 | 29.4 | 27.3 - 31.5 |
| HIV Status | Negative | 586 | 32.7 | 30.5 - 34.9 |
|  | Positive | 350 | 19.5 | 17.7 - 21.5 |
|  | Unknown | 856 | 47.8 | 45.4 - 50.1 |
| Residence Area | Rural | 263 | 14.7 | 13.1 - 16.4 |
|  | Urban | 1,529 | 85.3 | 83.6 - 86.9 |
| TB Treatment History | Previously treated | 339 | 18.9 | 17.1 - 20.8 |
|  | New | 785 | 43.8 | 41.5 - 46.1 |
|  | Unknown | 668 | 37.3 | 35 - 39.6 |
| GenXpert Result | Negative | 1,416 | 79.0 | 77 - 80.9 |
|  | TB-R | 18 | 1.0 | 0.6 - 1.6 |
|  | TB-S | 358 | 20.0 | 18.2 - 21.9 |
| TB Status | Drug Resistant * | 19 | 5.1 | 3.2 - 7.9 |
|  | Drug Susceptible | 357 | 94.9 | 92.1 - 96.8 |

\* A patient is defined as drug-resistant if he or she is resistant to at least one drug (RIF, INH, FQ, SLI).

Regarding HIV status, 32.7% (586/1792) of participants were HIV-negative, 19.5% (350/1792) were HIV-positive, while a significant proportion, 47.8% (856/1792), had an unknown HIV status. The high percentage of individuals with unknown HIV status represents a critical public health concern, given the well-established relationship between HIV infection and increased susceptibility to TB and its progression to active disease. This finding highlights a potential gap in integrated HIV testing and TB screening services within the healthcare system, underscoring the need for enhanced efforts to improve HIV diagnostic coverage among TB patients. Resistance profiles of mycobacterium tuberculosis samples is mentioned in Table2.

In terms of residence, the majority of participants resided in urban areas (85.3%, 1529/1792), while only 14.7% (263/1792) were from rural settings. The concentration of TB cases in urban centers likely reflects the higher population density, overcrowded living conditions, and potential barriers to timely healthcare access frequently encountered in urban informal settlements and slum areas. These environmental and socio-economic factors may facilitate the transmission of airborne diseases such as TB. Consequently, urban-centered TB control initiatives need to consider these underlying determinants to effectively reduce TB transmission rates.

Based on GeneXpert MTB/RIF assay results, 1.0% (18/1792) of participants tested positive for rifampicin-resistant TB (TB-R), 20.0% (359/1792) were rifampicin-sensitive (TB-S), and 79.0% (1415/1792) were negative for TB. The overall drug resistance rate was 5.1% (18/359) among those with confirmed TB, while 94.9% (341/359) were drug-susceptible.

The drug resistance profiles identified in our study are detailed in Table 2. The predominant resistance profile was multidrug-resistant TB (MDR-TB), defined as resistance to both rifampicin (RIF) and isoniazid (INH), observed in 94.44% (17/18) of the resistant isolates. This exceptionally high proportion of MDR-TB cases is concerning, as it complicates treatment management, requiring the use of longer, more expensive, and potentially toxic second-line drugs. This finding underscores the increasing threat of drug-resistant TB in the region.

**Table 2.** Resistance Profiles of Mycobacterium Tuberculosis samples.

|  | ***No of Isolates***<br>***(N=18)*** | ***Percentage***<br>***%*** |
| --- | --- | --- |
| RR | 7 | 38.88 % |
| INH | 1 | 5.55 % |
| RR+INH | 17 | 94.44 % |
| MDR+FQ | 2 | 11.11 % |
| MDR+SLI | 3 | 16.66 % |
| Total | 18 | 100 % |

Mono-resistance to rifampicin was detected in 38.88% (7/18) of resistant isolates. Although less common than MDR-TB, rifampicin resistance alone is a significant concern given its role as a cornerstone of standard first-line TB therapy. Resistance to isoniazid alone was less frequent, noted in 5.55% (1/18) of cases.

Furthermore, 11.11% (2/18) of resistant isolates exhibited MDR-TB with additional resistance to fluoroquinolones (MDR+FQ), and 16.66% (3/18) showed MDR with concurrent resistance to second-line injectable drugs (MDR+SLI). The emergence of these more advanced resistance profiles is worrisome, as it reduces effective treatment options and increases the risk of treatment failure and disease transmission. Notably, the presence of these pre-extensively drug-resistant (pre-XDR) cases signals the potential for further resistance evolution toward extensively drug-resistant TB (XDR-TB), emphasizing the urgency of reinforcing drug-resistance surveillance and programmatic management strategies.

Table 3 presents the analysis of factors associated with drug-resistant TB. Sex, age, residence area, and HIV status were not statistically significant risk factors for drug resistance. Female patients had an odds ratio (OR) of 0.61 (95% CI: 0.22–1.79, p=0.354) compared to males.

**Table 3.**
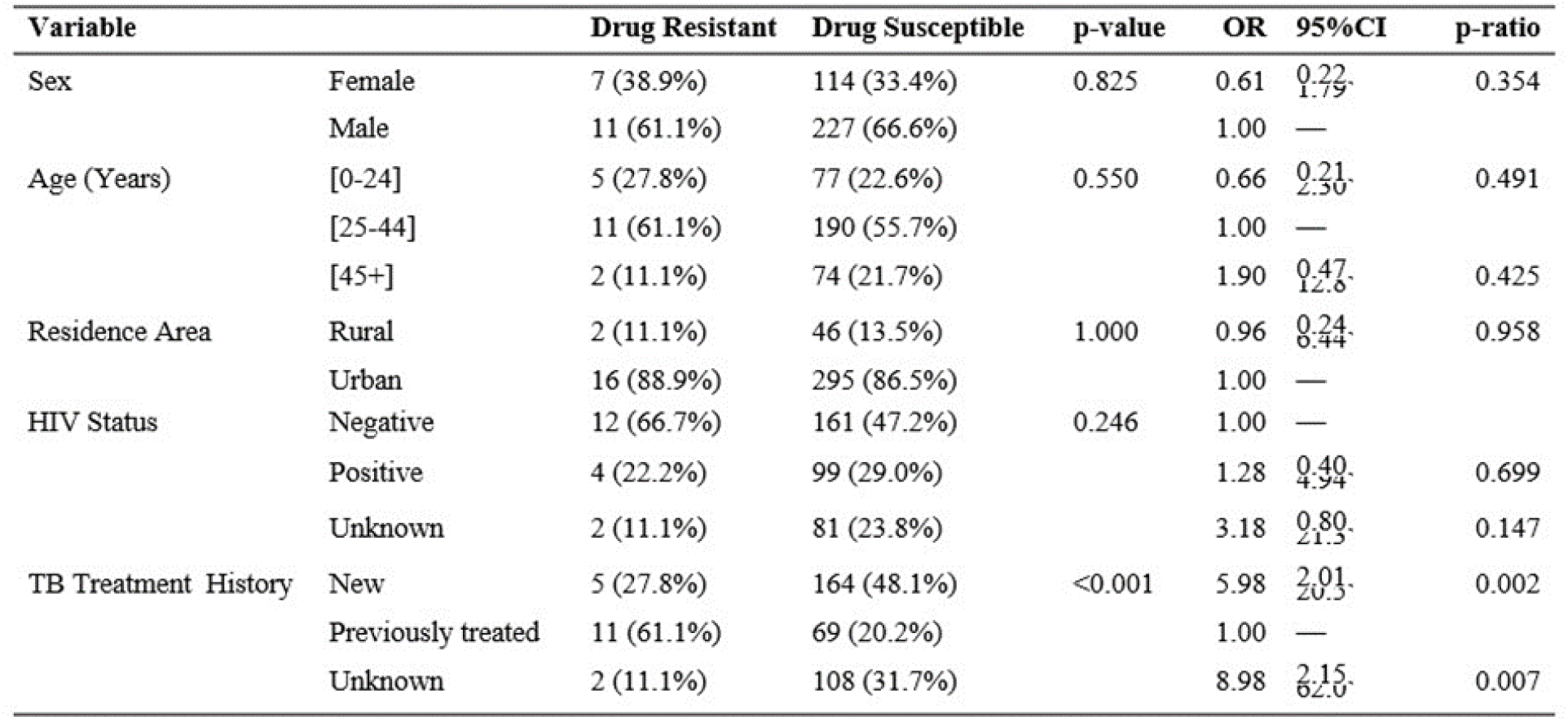
Factors associated with drug-resistant TB.

In contrast, previous TB treatment history was a significant predictor of drug resistance. Participants with a prior history of TB treatment had 3.18 times higher odds (95% CI: 1.08–9.35, p=0.037) of developing drug-resistant TB compared to new cases. Additionally, participants with unknown treatment histories exhibited an even higher risk (OR: 8.98, 95% CI: 2.15–37.50, p=0.007), likely reflecting unrecorded prior treatment exposures. These findings are consistent with existing literature and reaffirm the importance of comprehensive patient history documentation and adherence monitoring in TB control programs.

The most important risk factor for DMTB was its treatment history. Compared to new cases, cases previously treated showed marked increase in the risk for drug resistance (OR 3.18, 95% CI 1.08–9.35, p = 0.037). These findings hold true to the important role that prior treatment exposure as a risk factor for development of drug resistant TB.

However, demographic and characteristics of HIV related factors did not significantly affect RDR, buddied with prior TB treatment history that unswervingly sheathed record as a consistent and significant predictor yielding that full TB treatment documents and adherence to effective first line TB management protocols must be enforced to stem drug resistance in this setting.

## Discussion

This study provides valuable insights into the demographic characteristics of TB patients and the prevalence of drug-resistant TB in the Haut-Ogooué region. Several key findings warrant further discussion in the context of existing research and their implications for TB control efforts. The predominance of males among TB patients in our study, accounting for 62.1% of cases, is a recurring observation in numerous TB studies globally [1] . This pattern has been attributed to a complex interplay of factors. Social factors such as occupational exposures, health-seeking behavior, and smoking habits may contribute to the higher prevalence of TB in men. Biological factors, including hormonal differences and immune responses, may also play a role[7]. Understanding these gender disparities is crucial for designing targeted interventions to improve TB case detection and treatment adherence in men[7].

The age distribution of TB cases in our study, with the highest proportion in the 25-44 age group (37.8%), aligns with the findings of other studies that have reported a high burden of TB among individuals in their economically productive years [7]. This age group is often more mobile and engaged in social and occupational activities, increasing their risk of exposure to TB. The significant economic and social consequences of TB in this age group highlight the need for interventions that enable early diagnosis and prompt treatment to minimize transmission and maintain workforce productivity [8].

A noteworthy finding in our study is the high proportion of participants with unknown HIV status (47.8%). This is a significant concern, as HIV is a well-established risk factor for TB infection and progression to active disease . The lack of knowledge about HIV status among TB patients indicates a potential gap in HIV testing and counseling services. Integrated TB and HIV programs are essential for improving case detection, ensuring timely initiation of antiretroviral therapy for HIV-positive individuals, and ultimately reducing the burden of TB .

The concentration of TB cases in urban areas (85.5%) in our study underscores the role of urbanization in the spread of TB. Urban settings are often characterized by overcrowding, poverty, and inadequate housing, which create conducive conditions for TB transmission [9]. Rapid urbanization in many developing countries, including Gabon, poses a challenge to TB control efforts. Targeted interventions, such as improving housing conditions, reducing poverty, and strengthening healthcare services in urban slums, are crucial for addressing the TB burden in these settings [9].

The prevalence of multidrug-resistant TB (MDR-TB) in our study, with 94.44% of resistant isolates exhibiting resistance to both isoniazid and rifampicin, is a major concern. MDR-TB poses a significant threat to global TB control, as it requires the use of more toxic, less effective, and more expensive second-line drugs . The high proportion of MDR-TB in our study is consistent with reports from other settings in Africa, where drug resistance is an increasing problem. Factors such as inadequate treatment, poor adherence to treatment regimens, and weak health systems contribute to the emergence and spread of MDR-TB [10].

The detection of MDR-TB with resistance to second-line injectable drugs (16.66%) and pre-extensively drug-resistant TB (pre-XDR-TB) in our study is particularly alarming. Resistance to second-line drugs further limits treatment options and increases the risk of treatment failure [1]. The emergence of pre-XDR-TB highlights the urgent need to prevent the development of extensively drug-resistant TB (XDR-TB), which is even more difficult to treat [10].

Our study also showed a strong association between previous history of treating TB and drug-resistant TB. This was especially the case among patients who had previously been treated for TB. Such finding is consistent with a large body of evidence that points to returned treatment as a major risk factor for drug resistance. Factors, such as using a poor-quality drug, inadequate drug dosage, or poor adherence to treatment can lead to previous treatment selection, which can result in the selection of drug-resistant strains[10].

However, we did not observe a clear sex by drug resistant TB association as other studies have reported [10]. However, studies have found higher numbers of drug-resistant TB in men and our results indicate both men and women can be at risk. Additionally, we did not observe an important relation between age and drug-resistant TB, but there was a tendency towards elevated resistance among older patients.

Our study had a few limitations to bring to light. Some of the data collection was retrospective and this could’ve introduced bias. Furthermore, the study was limited to a single hospital, and so may not be generalized to other settings in Gabon.

## Limitations

Our study has a few limitations. Some data were collected retrospectively, which could introduce bias. It was also carried out at a single hospital, so the results may not reflect the situation in other parts of Gabon. Finally, we did not perform phenotypic drug susceptibility testing. Relying only on molecular tests means some resistance patterns might have been missed, and we could not compare genotypic results with the gold-standard culture-based method.

## Conclusion

This study provides critical insights into the landscape of TB and drug-resistant TB in the Haut-Ogooué region. A key factor associated with drug-resistant TB in our study population was a history of previous TB treatment. This finding aligns with existing research and emphasizes the importance of ensuring treatment adherence, providing adequate drug dosages, and utilizing quality-assured medications to prevent the selection and spread of drug-resistant strains. The concentration of TB cases in urban areas highlights the challenges posed by urbanization in TB control. Overcrowding and inadequate housing in urban settings likely contribute to TB transmission. In conclusion, our study emphasizes the high burden of MDR-TB and its association with previous TB treatment history. These findings indicate to strengthen TB control programs, improve treatment practices, and address the social determinants of TB in Gabon. Further research is needed to monitor drug resistance trends and evaluate the effectiveness of interventions aimed at curbing the spread of drug-resistant TB.

### What is already know on this topic

- Previous TB treatment history is a strong and consistent risk factor for developing drug-resistant TB, including MDR-TB.
- The majority of drug-resistant TB cases in the study were MDR-TB, accounting for 94.44% of resistant isolates.

### What this study adds

- A very high proportion of TB patients (47.8%) had unknown HIV status, indicating a major gap in integrated TB-HIV testing.
- Detection of pre-XDR TB (MDR-TB plus resistance to fluoroquinolones or injectables) suggests early warning signs of evolving extensively drug-resistant TB in the Haut-Ogooué region

## Data Availability

All data produced in the present study are available upon reasonable request to the authors

## Authors’ contributions

Conceptualization, **J.B.P.A.A.A**., **C.O.S**, ; and **M.E.D.N;** methodology, **P.S.E.O**; Data analysis, **M.E.D.N**., **J.B.P.A.A.A**., **P.S.E.O**., and **C.O.S**,.; validation, **J.B.P.A.A.A**., **M.E.D.N**., and **C.O.S**,.; Specimen collection, **P.S.E.O**., **T.E.; A.M**. and .; writing - original draft preparation, **J.B.P.A.A.A**., writing - review and editing **P.S.E.O** and **C.O.S**,; All authors have read and agreed to the published version of the manuscript.

## Acknowledgements (if any)

We are grateful to TB nurses and all the Laboratory staff for their hard work.

## References

1. World Health Organization. 2024 Global tuberculosis report. 2024.

2. Floyd K, Glaziou P, Houben RMGJ, Sumner T, White RG, Raviglione M. Global tuberculosis targets and milestones set for 2016–2035: definition and rationale. The International Journal of Tuberculosis and Lung Disease. 2018;22(7):723–730.

3. Hewison C, Ferlazzo G, Avaliani Z, Hayrapetyan A, Jonckheere S, Khaidarkhanova Z, et al. Six-Month Response to Delamanid Treatment in MDR TB Patients. Emerging Infectious Diseases. 2017;23(10):1746–1748.

4. Isaakidis P, Varghese B, Mansoor H, Cox HS, Ladomirska J, Saranchuk P, et al. Adverse Events among HIV/MDR-TB Co-Infected Patients Receiving Antiretroviral and Second Line Anti-TB Treatment in Mumbai, India. PLoS ONE. 2012;7(7):e40781.

5. Maier C, Chesov D, Schaub D, Kalsdorf B, Andres S, Friesen I, et al. Long-term treatment outcomes in patients with multidrug-resistant tuberculosis. Clinical Microbiology and Infection. 2023;29(6):751–757.

6. Jinyi W, Zhang Y, Wang K, Peng P. Global, regional, and national mortality of tuberculosis attributable to alcohol and tobacco from 1990 to 2019: A modelling study based on the Global Burden of Disease study 2019. Journal of Global Health. 2024;14. doi:10.7189/jogh.14.04023.

7. Murphy DA, Rotheram-Borus MJ, Joshi V. HIV-infected adolescent and adult perceptions of tuberculosis testing, knowledge and medication adherence in the USA. AIDS Care. 2000;12(1):59–63.

8. Grede N, Claros JM, de Pee S, Bloem M. Is There a Need to Mitigate the Social and Financial Consequences of Tuberculosis at the Individual and Household Level? AIDS and Behavior. 2014;18(S5):542–553.

9. Silva KRO, Ferreira RC, Coelho LE, Veloso VG, Grinsztejn B, Torres TS, et al. Knowledge of HIV transmission, prevention strategies and U = U among adult sexual and gender minorities in Brazil. Journal of the International AIDS Society. 2024;27(2). doi:10.1002/jia2.26220.

10. Seung KJ, Keshavjee S, Rich ML. Multidrug-Resistant Tuberculosis and Extensively Drug-Resistant Tuberculosis. Cold Spring Harbor Perspectives in Medicine. 2015;5(9):a017863.

